# Clinical predictors of severe forms of influenza A H1N1 in adults and children during the 2009 epidemic in Brazil

**DOI:** 10.1101/2023.09.08.23295269

**Authors:** Jose Ueleres Braga

## Abstract

The World Health Organization (WHO) raised the global alert level for the A(H1N1) influenza pandemic in June 2009. However, since the beginning of the epidemic, the fight against the epidemic lacked foundations for managing cases to reduce the disease lethality. It was urgent to carry out studies that would indicate a model for predicting severe forms of influenza. This study aimed to identify risk factors for severe forms during the 2009 influenza epidemic and develop a prediction model based on clinical epidemiological data. A case-control of cases notified to the health secretariats of the states of Rio de Janeiro, São Paulo, Minas Gerais, Paraná, and Rio Grande do Sul was conducted. Cases had fever, respiratory symptoms, positive confirmatory test for the presence of the virus associated with one of the three conditions: (i) presenting respiratory complications such as pneumonia, ventilatory failure, severe acute respiratory distress syndrome, sepsis, acute cardiovascular complications or death; or respiratory failure requiring invasive or non-invasive ventilatory support, (ii) having been hospitalized or (iii) having been admitted to an Intensive Care Unit. Controls were individuals diagnosed with the disease on the same date (or same week) as the cases. A total of 1653 individuals were included in the study, (858 cases/795 controls). These participants had a mean age of 26 years, a low level of education, and were mostly female. The most important predictors identified were systolic blood pressure in mmHg, respiratory rate in bpm, dehydration, obesity, pregnancy (in women), and vomiting (in children). Three clinical prediction models of severity were developed, for adults, adult women, and for children. The performance evaluation of these models indicated good predictive capacity. The area values under the ROC curve of these models were 0.89; 0.98 and 0.91 respectively for the model of adults, adult women, and children respectively.

## Introduction

In April 2009, the World Health Organization (WHO) communicated to all countries the occurrence of a public health emergency of international concern (April 2009) caused by infection with a new influenza A H1N1 virus. This infection was believed to be related to the febrile respiratory illness epidemic that had begun in Mexico in March 2009. The virus was finally detected in tests performed on two children in California, United States, on April 17(1).

This situation led the WHO to raise the pandemic alert level to level 5 and adopt recommendations aimed at reducing the effects of the infection based on the activation of national preparedness plans for the H5N1 influenza pandemic adapted to the new epidemic. The characteristics of this epidemic showed a predominance in children and young adults and a low lethality (<1%), with the majority of cases being of mild-to-moderate influenza-like illness and a higher frequency of deaths in patients with an underlying chronic disease; most cases recovered without specific treatment. Transmission occurred through direct contact or with the respiratory secretions of infected persons. The first estimates based on the analysis of cases in Mexico indicated a lower transmissibility than estimated for previous influenza pandemics (2) and for the severe acute respiratory syndrome (SARS).

Until June 2009, suspected cases of this disease in Brazil were linked to travel abroad or some type of close contact. However, in mid-July 2009, after concluding the epidemiological investigation of a suspected case in São Paulo, the country presented sustained transmission. This led to changes in the identification, investigation, and management of cases of flu syndrome, since any person who had flu symptoms would also be considered a suspected case of influenza A H1N1 infection. These changes were included in the “Influenza Clinical Management and Epidemiological Surveillance Protocol” (3). Following the recommendation of the World Health Organization (WHO), this scenario indicated the need to prioritize the notification, investigation, laboratory diagnosis, and treatment of cases with severe acute respiratory syndrome (SARS) and those people with risk factors for disease complications; these included individuals under 2 and over 60 years of age, pregnant women, those with chronic diseases, and immunosuppressed people. The new phase of studying the epidemic required monitoring cases in groups at risk of developing serious illness. At the time, Brazil was classified as a country that had a regional spread of this disease.

This study aims to identify the risk factors for the occurrence of severe forms during the 2009 influenza A H1N1 epidemic as well as to propose a prediction model based on clinical epidemiological data of cases notified to the Brazilian Epidemiological Surveillance System.

## Materials and Methods

The research used a case-control study design (4) of cases of influenza A H1N1 reported to the Health Surveillance of the health departments in the states of Rio de Janeiro, São Paulo, Minas Gerais, Paraná, and Rio Grande do Sul. The target population consisted of children, pregnant women, and adults with symptoms compatible with suspected infection by the influenza A H1N1 virus treated at the Health System.

Cases considered for this investigation were individuals with confirmed infection by the influenza A H1N1 virus with fever, respiratory symptoms, and a positive confirmatory test for the presence of the virus associated with one of the three conditions: (*i*) presenting respiratory complications such as pneumonia, ventilatory failure, severe acute respiratory distress syndrome, sepsis, acute cardiovascular complications, or death; or respiratory failure requiring invasive or non-invasive ventilatory support (CPAP, BIPAP, NIV, intubation)(4); (*ii*) having been hospitalized; or (iii) having been admitted to an intensive care unit.

To study the risk factors for the occurrence of severe cases of influenza A H1N1, three controls were chosen for each case. The criterion used was that they were individuals diagnosed with the disease on the same date (or in the same week) as the cases (5). Moreover, they should have been treated in the same health unit, hospital, or even intensive care unit if the case was treated there.

The minimum sample size was calculated for an alpha error of 5%, beta of 80%, and three controls for each case, resulting in 500 severe cases and 1,500 controls (6).

This study was approved by the Research Ethics Committee of the Institute of Social Medicine of Rio de Janeiro State University on December 4, 2009 (CAEE 0030.1.259.000- 09). It was an observational study with secondary data extracted from medical records in a standardized way in which access to the records respected the precepts of secrecy and confidentiality of data and had the consent of the person responsible for the health institution where the patient received care. So, the need for consent was waived by the ethics committee.

## Results

The research was based on data extracted from hospital records. It began in February 2010 with an initial review of the medical records of all patients with a confirmed diagnosis of influenza A H1N1 infection who were hospitalized in the selected cities.

This study included 1,653 individuals. The number of cases and controls distributed among the federative units were as follows: Paraná (253 cases and 154 controls), Rio Grande do Sul (73 cases and 67 controls), Minas Gerais (55 cases and 188 controls), São Paulo (387 cases and 111 controls), and Rio de Janeiro (90 cases and 275 controls).

These participants had a mean age of 26 years, and the study population ranged from newborns to older individuals. They had a low level of education and were mostly women. However, a large proportion of the sample did not reveal their education level, and no differences were observed in this profile when samples from the five federated units were compared. The proportion of pregnant women ranged from 8% to 43% of the samples, but the lack of information was between 30% and 60%; thus, the samples from the federated units showed homogeneity.

Regarding laboratory confirmation, it is worth mentioning that practically all the samples consisted of individuals who had given nasopharyngeal samples collected directly by swab. As for treatment outcomes, over 90% had favorable results, and mortality was 10% among those enrolled in this investigation.

Obesity was found in about 10% of patients, and it was the second most prevalent morbid condition. Only hypertension was more prevalent, affecting almost 14% of patients on average. Cystic fibrosis, tuberculosis, and rheumatoid arthritis were less prominent than the abovementioned diseases, and other less frequent morbid conditions included cerebrovascular disease, Lupus, and acute liver disease.

A small number of patients had some level of immunosuppression, and it included those who had undergone a transplant and were using corticosteroids. This group also comprised those with morbid conditions of intermediate prevalence such as chronic obstructive pulmonary disease (COPD) and diabetes, with proportions close to 5%. In addition, no significant variation was observed in the previous history of disease of the samples from the various federated units.

As for health care, one in every five patients was admitted to the ICU and had an average stay of nine days. More than a fifth of these patients were transferred from other units for hospitalization purposes.

The illness of the study population showed a highly diversified clinical picture. Cough and fever were the most prevalent signs and symptoms at the time of patient admission; convulsion, conjunctivitis, irritability, and drowsiness were the least frequent findings. More than half of the patients had dyspnea, while about a third had headaches, runny nose myalgia, and other symptoms.

Regarding the characteristics of the cases and controls classified by the severity condition adopted by the study, no significant differences were observed between the groups. The participants had similar age, gender, and level of education (Table 1).

**Table 1.** Biological and social characteristics, diagnostic conditions and outcomes of patients in the case-control study of risk factors for severity of H1N1 influenza, Brazil.

| Characteristic | Severity rating |  |  |
| --- | --- | --- | --- |
|  | severe | not severe | Total |
| N | 858 | 795 | 1653 |
| age (years) |  |  |  |
| average | 26.4 | 26.1 | 26.2 |
| Standart deviation | 18.8 | 18.9 | 18.9 |
| minimum | 0.0 | 0.1 | 0.0 |
| maximum | 90.0 | 94.0 | 94.0 |
| Sex (%) |  |  |  |
| female | 58.4 | 55.9 | 56.9 |
| male | 41.7 | 44.2 | 43.1 |
| Education (%) |  |  |  |
| Illiterate | 2.1 | 0.5 | 1.2 |
| 1st-4th incomplete series of PE | 3.5 | 4.5 | 4.1 |
| 4th complete series of PE | 1.2 | 0.7 | 0.9 |
| 5th-8th incomplete grades of PE | 3.7 | 3.5 | 3.6 |
| 8th grade complete PE or complete PE | 1.6 | 3.3 | 2.6 |
| Incomplete high school | 2.1 | 2.3 | 2.2 |
| Complete high school | 4 | 3.7 | 3.9 |
| incomplete higher education | 1.4 | 1.7 | 1.6 |
| complete higher education | 3.9 | 10.5 | 7.7 |
| Missing | 59.8 | 55.1 | 57.1 |
| Not applicable | 16.7 | 14.1 | 15.2 |
| Pregnance *(%) |  |  |  |
| yes | 26.8 | 23.3 | 24.9 |
| no | 32 | 32.9 | 32.5 |
| missing | 41.2 | 43.8 | 42.6 |
| Condition of laboratory confirmation (%) |  |  |  |
| live | 96.7 | 95.1 | 96.0 |
| post mortem | 0.4 | 1.1 | 0.7 |
| missing | 2.9 | 3.9 | 3.3 |
| Laboratory confirmation material (%) |  |  |  |
| nasopharyngeal swab | 95.1 | 96.7 | 96.0 |
| tissue fragments | 1.1 | 0.4 | 0.7 |
| missing | 3.9 | 2.9 | 3.3 |
| Disease outcome (%) |  |  |  |
| death | 20.0 | 4.0 | 10.9 |
| discharge/cure | 78.9 | 94.8 | 88.0 |
| hospital transfer | 1.1 | 0.7 | 0.8 |
| missing | 0.0 | 0.5 | 0.3 |

Laboratory conditions for confirming the disease in H1N1 cases and controls were similar. However, patient outcomes differed between these groups, and the proportion of deaths was five times greater among cases than controls, which is a statistically significant difference. The other characteristics were similar (Table 1).

Obesity, which was prevalent in 10% of patients, and hypertension were similar between cases and controls. The other morbid conditions studied were similar in both groups. Less frequent morbid conditions that were similar among participants in both groups were cystic fibrosis, tuberculosis, and rheumatoid arthritis as well as cerebrovascular diseases, Lupus, and acute liver diseases (Table 2). Other past diseases also had similar distributions in the studied groups.

**Table 2.** History of previous illnesses of cases and controls of the case-control study of risk factors for severity of H1N1 influenza, Brazil.

| Condition | Severity rating |  |  | Condition | Severity rating |  |  |
| --- | --- | --- | --- | --- | --- | --- | --- |
|  | severe | not severe | total |  | severe | not severe | total |
| N | 407 | 498 | 1653 | N | 407 | 498 | 1653 |
| Obesity (%) |  |  |  | Acute liver disease (%) |  |  |  |
| yes | 11.3 | 10.0 | 10.5 | yes | 0.5 | 0.4 | 0.5 |
| no | 88.8 | 90.0 | 89.5 | no | 99.5 | 99.6 | 99.6 |
| Hypertension (%) |  |  |  | Chronic liver disease (%) |  |  |  |
| yes | 14.9 | 12.9 | 13.8 | yes | 1.4 | 0.9 | 1.1 |
| no | 85.1 | 87.1 | 86.2 | no | 98.6 | 99.1 | 98.9 |
| Coronary Disease (%) |  |  |  | Hematological Disease (%) |  |  |  |
| yes | 2.5 | 2.7 | 2.6 | yes | 4.4 | 4.1 | 4.2 |
| no | 97.5 | 97.3 | 97.4 | no | 95.6 | 95.9 | 95.8 |
| CVD (%) |  |  |  | Lupus (%) |  |  |  |
| yes | 0.4 | 0.8 | 0.6 | yes | 0.5 | 0.7 | 0.6 |
| no | 99.7 | 99.2 | 99.4 | no | 99.5 | 99.3 | 99.4 |
| Heart Failure (%) |  |  |  | Rheumatoid arthritis (%) |  |  |  |
| yes | 2.5 | 1.5 | 1.9 | yes | 0.4 | 0.1 | 0.2 |
| no | 97.5 | 98.5 | 98.1 | no | 99.7 | 99.9 | 99.8 |
| Diabetes (%) |  |  |  | HIV (%) |  |  |  |
| yes | 6.3 | 5.9 | 6.1 | yes | 1.6 | 2 | 1.8 |
| no | 93.7 | 94.2 | 93.9 | no | 98.4 | 98 | 98.2 |
| Thyroid disease (%) |  |  |  | AIDS (%) |  |  |  |
| yes | 3.2 | 3.1 | 3.1 | yes | 1.1 | 1.5 | 1.3 |
| no | 96.8 | 96.9 | 96.9 | no | 99 | 98.5 | 98.7 |
| COPD (%) |  |  |  | Neoplasm (%) |  |  |  |
| yes | 5.3 | 3.5 | 4.5 | yes | 2.3 | 2.8 | 2.6 |
| no | 94.7 | 96.5 | 95.5 | no | 97.7 | 97.2 | 97.4 |
| Tuberculosis (%) |  |  |  | Organ transplantation (%) |  |  |  |
| yes | 0.0 | 0.1 | 0.1 | yes | 0.7 | 1.1 | 0.9 |
| no | 100 | 99.9 | 99.9 | no | 99.3 | 98.9 | 99.1 |
| Cystic fibrosis (%) |  |  |  | Use of Corticosteroids (%) |  |  |  |
| yes | 0.0 | 0.1 | 0.1 | yes | 3.5 | 5.2 | 4.5 |
| no | 100 | 99.9 | 99.9 | no | 96.5 | 94.8 | 95.5 |

Regarding the history of assistance received, the groups showed important differences. For the presence of signs and symptoms upon hospital admission, severe cases had more important respiratory symptoms, such as dyspnea and cough, compared to controls; they also showed a greater occurrence of fever. The other symptoms did not show a higher frequency among severe cases. In fact, for less specific clinical symptoms, these patients had a highly similar profile (Table 3).

**Table 3.** Symptoms reported on hospital admission of cases and controls from the case-control study of risk factors for severity of H1N1 influenza, Brazil.

| Condition | Severity rating |  |  | Condition | Severity rating |  |  |
| --- | --- | --- | --- | --- | --- | --- | --- |
|  | severe | not severe | total |  | severe | not severe | total |
| N | 407 | 498 | 1653 |  | 407 | 498 | 1653 |
| Fever (%) |  |  |  | Nasal congestion (%) |  |  |  |
| yes | 96.5 | 91.9 | 93.9 | yes | 12.5 | 12.8 | 12.6 |
| no | 2.1 | 5.3 | 3.9 | no | 21.4 | 32.3 | 27.6 |
| missing | 1.4 | 2.8 | 2.2 | missing | 66.1 | 54.9 | 59.7 |
| Cough (%) |  |  |  | Diarrhea (%) |  |  |  |
| yes | 94.4 | 91 | 92.4 | yes | 10.4 | 9.3 | 9.8 |
| no | 0.7 | 3.1 | 2.0 | no | 33.9 | 43.9 | 39.6 |
| missing | 4.9 | 6.0 | 5.5 | missing | 55.7 | 46.8 | 50.6 |
| Headache (%) |  |  |  | Vomiting (%) |  |  |  |
| yes | 25.7 | 35.1 | 31.0 | yes | 19.5 | 19.0 | 19.2 |
| no | 17.6 | 24.6 | 21.6 | no | 26.7 | 36.2 | 32.1 |
| missing | 56.8 | 40.3 | 47.4 | missing | 53.8 | 44.8 | 48.7 |
| Chill (%) |  |  |  | Seizures (%) |  |  |  |
| yes | 12.5 | 9.2 | 10.6 | yes | 1.6 | 2.1 | 1.9 |
| no | 25.1 | 35.5 | 31.0 | no | 25.7 | 41.9 | 34.9 |
| missing | 62.4 | 55.3 | 58.4 | missing | 72.8 | 56.0 | 63.2 |
| Dyspnea (%) |  |  |  | Asthenia (%) |  |  |  |
| yes | 75.2 | 54.5 | 63.4 | yes | 22.1 | 21.9 | 22 |
| no | 9.1 | 23.3 | 17.2 | no | 14.8 | 27.0 | 21.7 |
| missing | 15.6 | 22.2 | 19.4 | missing | 63.1 | 51.1 | 56.3 |
| Sore throat (%) |  |  |  | Lack of appetite (%) |  |  |  |
| yes | 18.3 | 25.7 | 22.5 | yes | 18.1 | 15.6 | 16.7 |
| no | 26.2 | 31.7 | 29.3 | no | 18.6 | 30.3 | 25.3 |
| missing | 55.5 | 42.7 | 48.2 | missing | 63.3 | 54.1 | 58.1 |
| Arthralgia (%) |  |  |  | Irritability (%) |  |  |  |
| yes | 10 | 7.7 | 8.7 | yes | 3.3 | 2.0 | 2.6 |
| no | 27.9 | 37.5 | 33.4 | no | 22.5 | 37.1 | 30.8 |
| missing | 62 | 54.8 | 57.9 | missing | 74.2 | 60.9 | 66.6 |
| Myalgia (%) |  |  |  | Somnolence (%) |  |  |  |
| yes | 45.5 | 43.9 | 44.6 | yes | 5.3 | 3.2 | 4.1 |
| no | 13.5 | 22.5 | 18.6 | no | 23.7 | 37.9 | 31.8 |
| missing | 41 | 33.6 | 36.8 | missing | 71 | 58.9 | 64.1 |
| Conjunctivitis (%) |  |  |  | Other symptoms (%) |  |  |  |
| yes | 1.6 | 2.4 | 2.0 | yes | 47.1 | 32.5 | 38.8 |
| no | 31.8 | 41.1 | 37.1 | no | 23.4 | 47.5 | 37.1 |
| missing | 66.6 | 56.5 | 60.9 | missing | 29.5 | 20.1 | 24.2 |
| Runny nose(%) |  |  |  |  |  |  |  |
| yes | 30.4 | 36.8 | 34.1 |  |  |  |  |
| no | 20.7 | 26.7 | 24.2 |  |  |  |  |
| missing | 48.9 | 36.4 | 41.8 |  |  |  |  |

Initially, exploratory analyses were conducted to identify the main factors related to the severity of cases of influenza A H1N1. The factors described in the literature and those aspects whose frequency of evaluation and registration in the medical records allowed for analysis were taken as a basis. The first model studied and constructed was the one related to the severity of cases in adults (Table 4).

**Table 4.** Risk factors associated with the severity of influenza A (H1N1) in adults, Brazil.

| Risk factor | crude OR | CI 95% | adj OR | CI 95% |
| --- | --- | --- | --- | --- |
| **Adult** |  |  |  |  |
| Systolic Blood Pressure in mmHg | 1.01 | 1.01 - 1.02 | **1.02** | **1.01 - 1.03** |
| Diastolic Blood Pressure in mmHg | 1.00 | 0.99 - 1.01 | 0.99 | 0.96 - 1.01 |
| Heart Rate in bpm | 1.00 | 0.99 - 1.01 | 0.99 | 0.97 - 1.01 |
| Respiratory Rate in bpm | 1.06 | 1.05 - 1.07 | **1.05** | **1.01 - 1.08** |
| Axillary temperature in °C | 1.01 | 0.89 - 1.13 | 1.10 | 0.90 - 1.34 |
| Mental confusion | 3.56 | 1.55 - 8.21 | 1.40 | 0.31 - 6.20 |
| Dehydration | 2.20 | 1.44 - 3.39 | **2.69** | **1.34 - 5.39** |
| Impaired general condition/Toxemia/Prostration | 2.62 | 1.93 - 3.57 | 1.29 | 0.73 - 2.26 |
| Cyanosis | 2.69 | 1.44 - 4.99 | 1.01 | 0.29 - 3.39 |
| Somnolence | 4.25 | 1.85 - 9.76 | 18.95 | 0.45 - 7.97 |
| Obesity | 1.68 | 1.08 - 2.61 | 1.60 | 0.81 - 3.19 |
| **Adult (women only)** |  |  |  |  |
| Systolic Blood Pressure in mmHg | 1.01 | 1.01 - 1.02 | 1.03 | 0.98 - 1.08 |
| Diastolic Blood Pressure in mmHg | 1.01 | 0.99 - 1.02 | 0.96 | 0.89 - 1.05 |
| Heart Rate in bpm | 1.01 | 0.99 - 1.02 | 0.97 | 0.94 - 1.01 |
| Respiratory Rate in bpm | 1.05 | 1.00 - 1.08 | 1.04 | 0.97 - 1.12 |
| Axillary temperature in °C | 1.04 | 0.88 - 1.22 | 0.96 | 0.50 - 1.82 |
| Mental confusion | 3.54 | 1.24 - 10.11 |  |  |
| Dehydration | 1.94 | 1.10 - 3.42 | **30.36** | **2.85 - 3.23** |
| Impaired general condition/Toxemia/Prostration | 2.28 | 1.55 - 3.37 | 0.95 | 0.23 - 3.94 |
| Cyanosis | 1.77 | 0.82 - 3.82 |  |  |
| Somnolence | 3.43 | 1.17 - 10.09 | 4.20 | 0.19 - 9.22 |
| Obesity | 1.94 | 1.09 - 3.45 | **3.84** | **1.06 - 13.83** |
| Pregnancy | 1.15 | 0.78 - 1.70 | **5.43** | **1.27 - 23.13** |
| **Child** |  |  |  |  |
| Systolic Blood Pressure in mmHg | 1.00 | 0.98 - 1.02 | 0.99 | 0.96 - 1.04 |
| Diastolic Blood Pressure in mmHg | 1.01 | 0.99 - 1.04 | 1.04 | 0.98 - 1.10 |
| Heart Rate in bpm | 1.02 | 1.01 - 1.03 | 1.01 | 0.99 - 1.04 |
| Respiratory Rate in bpm | 1.06 | 1.04 - 1.08 | **1.06** | **1.01 - 1.12** |
| Axillary temperature in °C | 0.99 | 0.91 - 1.08 | 0.82 | 0.46 - 1.44 |
| Nose wing beat | 1.23 | 1.15 - 1.32 | 0.93 | 0.68 - 1.27 |
| Intercostal indrawing | 1.17 | 1.09 - 1.25 | 1.20 | 0.89 - 1.60 |
| Dehydration | 0.98 | 0.89 - 1.07 | 1.17 | 0.88 - 1.57 |
| Impaired general condition/Toxemia/Prostration | 1.02 | 0.94 - 1.11 | 0.89 | 0.71 - 1.13 |
| Cyanosis | 1.03 | 0.95 - 1.12 | 0.80 | 0.63 - 1.03 |
| Lack of appetite | 1.12 | 1.05 - 1.20 | 0.82 | 0.65 - 1.05 |
| Vomiting | 1.20 | 1.12 - 1.28 | **1.32** | **1.08 - 1.61** |
| Difficulty swallowing fluids | 1.18 | 1.10 - 1.27 | 1.14 | 0.87 - 1.48 |
| Somnolence | 1.16 | 1.09 - 1.25 | 1.16 | 0.93 - 1.46 |

Considering only adult patients, the factors most strongly associated with severity in the bivariate analysis were drowsiness and mental confusion. Only the signs of systolic blood pressure and respiratory rate were associated with statistical significance at the 5% level. Obesity was associated with severity when putative confounders were not considered.

After adjustment for the set of clinical factors, it was observed that only systolic blood pressure, respiratory rate, and dehydration remained associated when multiple logistic regression was used. In addition to these aspects, biological factors such as age and sex were evaluated and were not associated with severity (Table 4).

As pregnancy is considered a condition that may be associated with severity, it was necessary to make a specific model for women. In this case, in addition to the aspects used for the general adult population, the condition of pregnancy was included among the factors studied (Table 4).

Similar to the prediction model studied for adults in general, in pregnant women, most of the factors described in the literature were associated with severity in the bivariate context, which draws attention to the importance of dehydration appearing in this adult model. However, the aspects that should be emphasized the most are obesity, which gains importance for predicting severity, and pregnancy, which appears to be the second most important factor for predicting this clinical condition (Table 4).

As we considered the elaboration of a specific model for women, we also decided to construct a specific one for the group of patients under 13 years of age as the factors that influence the occurrence of severity in this group are different from those affecting adults.

The specific literature on risk factors for severe forms of influenza A H1N1 indicates a set of factors seemingly more specific to this age group, such as the existence of clinical signs of intercostal retraction and nasal flaring, in addition to classic ones such as dehydration.

Similar to the models built for predicting severity in adults, exploratory analyses were initially performed to identify important associations with all the factors described in the literature, using data extracted from the medical records.

In patients younger than 13 years, despite the large number of factors associated with the severity of influenza A H1N1 in the bivariate context, no strong associations were found with the factors observed in adult models. Biological aspects such as age and gender were not associated, even when considered without adjustment for the other factors; the same occurred for obesity. It is noteworthy that, as in the adult model, only respiratory rate and the occurrence of vomiting were associated with severe disease in the multivariate context (Table 4).

The models built in this work show good performance as they included performance measures that indicate good predictive capacity. The area under the ROC curve values of these models were 0.89, 0.98, and 0.91 for the model of adults, adult women, and children, respectively.

## Discussion

The main research findings were the prediction models of severe forms of influenza A H1N1 specific for adults, adult women, and children. These models showed a high ability to discriminate severe forms since they have high values of area under the curve. However, the fact that these models were produced based on the occurrence of the 2009 influenza epidemic in Brazil could bring limitations about the external validity of this prediction capacity (7). The most important predictors identified in our research were systolic blood pressure in mmHg, respiratory rate in bpm, dehydration, obesity, pregnancy (in women), and vomiting (in children). It is worth noting that the good performance of these models can help improve approaches to surveillance, case management, and coping with future epidemics.

The need to overcome the difficulties of constructing and validating prediction models to predict the most severe clinical conditions during the epidemic has mobilized several researchers to disseminate epidemiological methods applied to the construction and validation of these models (8). Even so, the absence of external validation in the development process does not make models less important than those produced in our work.

The literature shows few studies of construction and validation of prediction related to influenza. Two stand out—a study on the prediction of respiratory infection by the influenza virus (9) and a systematic review of models for predicting this viral infection (10). However, no publication is available on the development and validation of a prediction model for severe forms including adults, adult women, and children.

Concerning risk factors for severe forms of influenza A H1N1, it should be noted that few studies have been published in the literature. Many studies have described the characteristics of severe cases and reported that underlying chronic diseases, delay in antiviral therapy, pregnancy, and obesity may be risk factors (11)(12)(13).

A study conducted in the United Kingdom (14) that used an epidemiological design (case note-based investigation) whose control was the population of London and Northern Ireland (> 70% of cases) sought to identify risk factors for hospitalization and outcomes unfavorable conditions during the first wave of the 2009 influenza A H1N1 pandemic. The authors concluded that an abnormal chest X-ray or an elevated CRP level indicates a potentially serious outcome, especially in patients registered as obese or who have lung diseases other than asthma or COPD. They also indicated that these findings support the use of the pandemic vaccine in pregnant women, children < 5 years of age, and people with chronic lung disease. Similarly, Myles et al. (15) sought to identify predictors of clinical outcomes in a national cohort hospitalized during both waves of the 2009–2010 influenza A H1N1 epidemic in the United Kingdom, using a profile comparison of patients with seasonal winter admissions by acute respiratory infection. They concluded that independent predictors of severe outcome were age between 55 and 64 years, chronic lung disease (not asthma, non-chronic obstructive pulmonary disease), neurological disease, registered obesity, late admission (≥ 5 days after illness onset), pneumonia, C-reactive protein ≥ 100 mg/liter, and the need for supplemental oxygen or intravenous fluid replacement on admission.

Fang et al. (16) conducted a case-control study to explore risk factors for severe cases of the 2009 influenza A H1N1, with mild cases as controls. They randomly selected mild and severe cases (230 cases each) from nine cities in Zhejiang Province, China, and performed a case-control study. They found that it took an average of five days for severe 2009 influenza A H1N1 cases to start antiviral therapy far from baseline—two days after mild cases. Having underlying chronic illnesses and poor psychological health combined with underlying chronic illnesses were two important risk factors for severe cases, and timely antiviral therapy was a protective factor. The authors concluded that psychological health education and intervention, as well as timely antiviral therapy, could not be ignored in the prevention, control, and treatment of 2009 influenza A H1N1.

Ren et al. (17) conducted a case-control study with 343 critically ill hospitalized patients and 343 randomly selected mild controls. The diagnosis was established by assessing clinical symptoms and confirmed by real-time reverse transcriptase polymerase chain reaction assay. Severe or mild patients were classified according to uniform criteria issued by the Chinese Ministry of Health. According to the authors, multivariate logistic regression analysis showed that overweight or obese individuals admitted to the hospital with H1N1 flu were more likely to have severe manifestations of the disease. Moreover, individuals younger than 5 years old or older than 60 years old had an increased risk of severe manifestations. They also observed an increased risk among individuals with a longer time interval from the onset of symptoms to hospital admission and found that those with chronic illnesses were at increased risk of severe manifestations of the H1N1 flu.

Studies on the risk factors for hospitalization for influenza A H1N1 have been conducted in several countries, finding the causes of hospitalization to be many stools as a result of severe evolution of clinical conditions. Launes et al. (18) conducted a multicenter study (case-control design) to compare sociodemographic data and preexisting medical conditions of risk in patients with 2009 influenza A H1N1 infection who required hospitalization and those treated on an outpatient basis. Cases were patients aged 6 months to 18 years hospitalized for influenza syndrome, for whom infection with the 2009 influenza A H1N1 was confirmed using transcription polymerse chain reaction (TR-PCR). Controls were patients aged between 6 months and 18 years with confirmed 2009 influenza A H1N1, treated as outpatients. Cases were 195 and controls 184. Launes et al. concluded that hospitalization was more frequent in children aged < 2 years, in those with neurological and/or neuromuscular diseases, and those whose parents had an education level lower than high school. Children younger than 2 years old, with neurological diseases, and from families with a lower level of education were at higher risk of hospitalization due to infection with 2009 influenza A H1N1.

Another study of risk factors for hospitalization was conducted by Ward et al. (19) in Sydney, Australia, which included included 302 cases and 603 controls. In a logistic regression model, after adjusting for age and sex, risk factors for hospitalization were pregnancy, immunosuppression, pre-existing lung disease, asthma requiring regular preventive medication, heart disease, diabetes, and current smoker or former smoker.

Some investigations have sought to answer whether obesity is a predictor of the evolution of influenza cases that occur during an epidemic. A case-control study was conducted in seven regions of Spain to investigate the association between body mass index (BMI) and the risk of hospitalization for influenza in 2009–2010 and 2010–2011. Martin et al. (20) selected cases among patients hospitalized for > 24 h with influenza-like illness, acute respiratory infection, septic shock, or multiple organ failure among whom 2009 influenza A H1N1 virus infection was confirmed by reaction in polymerase chain with real-time reverse transcription (RT-PCR). Hospitalized patients (n = 755) with laboratory-confirmed influenza were individually matched by age, date of admission/appointment, and province, with an outpatient (n = 783) with laboratory-confirmed influenza and an outpatient control (n = 950). Higher BMI was associated with an increased risk of hospitalization compared with outpatient cases and outpatient controls. Compared with normal weight, type I obesity, type II obesity, and type III obesity were associated with a higher likelihood of hospitalization compared with outpatient cases. Compared with normal weight, type II obesity and type III obesity were associated with a higher likelihood of hospitalization compared with outpatient controls.

A condition not investigated in our study was whether the vaccination status for the specific influenza vaccine would be a predictor of severe forms of the disease. This was not possible because no vaccination was conducted in Brazil during the investigation’s field phase. However, enough studies are available for systematic reviews with meta-analyses to be produced. The systematic review conducted by Rondy et al. (21) included case-control studies; the authors identified 3,411 publications, of which 30 met the inclusion criteria. Between 2010–11 and 2014–15, vaccine effectiveness was 41% (95% CI: 34; 48) for any flu (51%–95% CI: 44; 58).

We consider that the three prediction models of severe clinical forms of influenza A H1N1 specific for adults, adult women, and children can be used to address future influenza epidemics—at least to guide the investigation of cases with a greater probability of evolving into severe forms.

## Data Availability

Data cannot be shared publicly because of Data are not available because the authors have committed themselves to secrecy and confidentiality in accordance with the agreement of the Brazilian Research Ethics Committee

## Acknowledgments

We appreciate the help of collaborators Ana Freitas Ribeiro, Guilherme Loureiro Werneck, Bernardino Claudio de Albuquerque, Sonia Raboni and, Ricardo Kuchenbecker.

